# Optimizing Prescribing in Older Adults with Multimorbidity and Polypharmacy in Primary Care: A Cluster Randomized Clinical Trial (OPTICA Trial)

**DOI:** 10.1101/2022.10.31.22281164

**Authors:** Katharina Tabea Jungo, Anna-Katharina Ansorg, Carmen Floriani, Zsofia Rozsnyai, Nathalie Schwab, Rahel Meier, Fabio Valeri, Odile Stalder, Andreas Limacher, Claudio Schneider, Michael Bagattini, Sven Trelle, Marco Spruit, Matthias Schwenkglenks, Nicolas Rodondi, Sven Streit

## Abstract

**Importance:** Inappropriate prescribing and prescribing omissions are major drivers of healthcare-related harm. Medication review may help improve pharmacotherapy.

**Objective:** To study the effects of a primary care medication review intervention centered around an electronic clinical decision support system (eCDSS) on medication appropriateness and the number of prescribing omissions in older adults with multimorbidity and polypharmacy compared to usual care.

**Design and Setting:** The “Optimising PharmacoTherapy In the multimorbid elderly in primary Care” (OPTICA) trial is a cluster randomized clinical trial conducted with general practitioners (GPs) and older multimorbid patients with polypharmacy in Swiss primary care settings, between January 2019 and February 2020. The 12-month follow-up was completed in February 2021.

**Participants:** Eligible patients had to be ≥65 years of age with ≥3 chronic conditions and ≥5 long-term medications.

**Intervention:** The intervention to optimize pharmacotherapy centered around an eCDSS compared to usual care.

**Main Outcomes and Measures:** The two primary outcomes were the improvement in the Medication Appropriateness Index (MAI) and the Assessment of Underutilization (AOU) at 12 months. Secondary outcomes included the number of medications, number of falls and fractures and quality of life.

**Results:** In 43 GP clusters, 323 patients were recruited (median age: 77 years (IQR: 73-83), 45% were female). 21 GPs with 160 patients were assigned to the intervention group and 22 GPs with 163 patients to the control group. On average, 1 recommendation to stop or start a medication were reported to be implemented per patient. At 12 months, there were no group differences in the improvement of medication appropriateness (Odds ratio (OR): 1.05; 95% confidence interval (CI): 0.59 to 1.87) nor the number of prescribing omissions (OR: 0.90; 95% CI: 0.41 to 1.96) in the intention-to-treat analysis. The per-protocol analysis showed no statistically significant group difference and there were no group differences in the secondary outcomes either.

**Conclusions and Relevance:** In this randomized trial of GPs and older adults, medication reviews based on the eCDSS reduced inappropriate prescriptions but did not lead to higher appropriateness of patients’ medications. The intervention could be safely delivered to patients without causing any detriment to their health.

**Funding:** Swiss National Science Foundation (407440_167465)

**Key points:** 

**Question:** What is the effect of a GP administered medication review intervention supported by an electronic clinical decision support system on medication appropriateness in older patients with multimorbidity and polypharmacy?

**Findings:** This cluster randomized controlled trial included 43 general practitioners and 323 patients. At the end of the 12-month follow-up period, medication appropriateness and the number of prescribing omissions did not differ between patients who received the intervention and those who received usual care.

**Meaning:** The intervention to optimize pharmacotherapy was feasible and safe to implement in primary care but did not improve overall medication appropriateness nor reduce the number of prescribing omissions.

## Introduction

Inappropriate polypharmacy is highly prevalent in older adults and a major driver of healthcare-related harm.^1,2^ It is associated with negative health outcomes, such as adverse drug events, falls and functional decline in activities of daily living.^3-6^ Patients with multiple chronic conditions (multimorbidity^7^) and polypharmacy, defined as the use of ≥5 medications,^8^ are at an increased risk of inappropriate polypharmacy, such as inappropriate prescribing^9-11^ and prescribing omissions.^12^ This highlights the need for reducing inappropriate polypharmacy. Primary care settings, characterized by long-term patient-provider relationships, lend itself as ideal settings for medication reviews. Performing medication reviews, however, is a complex and time-consuming task. And currently, the evidence for medication review interventions is mixed.^13-15^

Medication review interventions, such as those based on the ‘Screening Tool of Older Persons’ potentially inappropriate Prescription’ (STOPP) and ‘Screening Tool to Alert doctors to the Right Treatment’ (START) criteria – an evidence-based criteria to inform prescribing in older adults^16,17^– can support physicians. These criteria have been shown to be effective in improving prescribing quality and some patient outcomes.^18,19^ In the context of digitalization, it is a promising way forward to use electronic clinical decision support systems (eCDSS) based on the STOPP/START criteria, such as the ‘Systematic Tool to Reduce Inappropriate Prescribing’-Assistant (STRIPA).^20-22^ In recent years, different eCDSS for optimizing medication use have been tested.^23-25^ Systems based on the STOPP/START criteria were used in two randomized clinical trials conducted in the inpatient setting.^19,26^ However, evidence from primary care settings is lacking.

The OPTICA trial tested the hypothesis that in older adults with multimorbidity and polypharmacy the use of an eCDSS for optimizing pharmacotherapy by general practitioners (GPs) improves medication appropriateness and reduces prescribing omissions more than standard care.

## Methods

### Trial design

The protocol of the ‘Optimising PharmacoTherapy In the multimorbid elderly in primary CAre’ (OPTICA) trial was published previously.^27^ We conducted a cluster randomized controlled trial with 43 GPs as clusters. GPs recruited 8-10 of their older patients with multimorbidity and polypharmacy. The trial, conducted from 2018 to 2021, was approved by the responsible ethics committee before enrolment commenced (BASEC ID: 2018–00914). We obtained written informed consent from all participants or their legal representatives prior to study enrolment.

### Participants

GPs had to be participating in the *‘Family medicine ICPC Research using Electronic medical records*’ (FIRE) project.^28^ This allowed electronic health record (EHR) data exports from their GP practices to the trial database and the eCDSS.

Patients were ≥65 years of age, to take ≥5 long-term medications (≥90 days) and to have ≥3 chronic conditions based on ICPC-2 coding or GPs’ clinical judgement. In case eligible patients suffered from cognitive impairment, written informed consent was obtained from their legal representative.

### Randomization

Randomization of clusters was done after patient enrolment for each cluster was completed. Randomization of participating GPs was done centrally in a web-based system (REDCap).^29,30^ We used a 1:1 ratio with unstratified block randomization and randomly varying block sizes of two and four.

### Trial procedures

*Intervention group:* The intervention was performed by the GPs at the individual patient level and consisted of a structured medication review using STRIPA, a web-based electronic clinical decision support system based on the ‘Screening Tool of Older Person’s Prescriptions’ (STOPP) and ‘Screening Tool to Alert doctors to Right Treatment’ (START) criteria V.2.^17,31^ In addition to detecting potential drug overuse, underuse, and misuse, STRIPA generated recommendations to address drug-drug interactions and adapt the dosage of inappropriate prescriptions.

*Control group:* GPs were asked to discuss their patients’ medications in accordance with their usual practice.

### Blinding

GPs were blinded during the screening and recruitment process of patients to limit biased selection of patients. GPs in the control group remained partially blinded, as they did not know the intervention procedure. Patients in the control group received a sham intervention (medication discussion). The data collectors and study assessors were fully blinded. Blinding of the trial statistician was not feasible because the data export contained information on the study groups in various forms.

### Outcomes

#### Primary outcome measures

Medication appropriateness was the primary outcome. To account for the multi-dimensionality of this construct, we used two primary outcome measures: the Medication Appropriateness Index (MAI) and the Assessment of Underutilization (AOU) (both at 12 months). The MAI helps assess the appropriateness of prescriptions and the AOU is a tool to evaluate under-prescribing by measuring the number of prescribing omissions.^32-34^ We assessed the AOU for each non-acute condition of the patients and the MAI for each long-term medication. We used the 10-item version of the MAI. However, we excluded the cost-effectiveness item for feasibility reasons (**Appendix 1**). This resulted in a score from 0 to 17 for each medication (with a higher score representing greater inappropriateness). Improvement in the MAI was defined as a decrease of ≥1 score point. For the AOU, we assessed for every non-acute diagnosis whether there was i) no prescribing omission, ii) a marginal omission (e.g. use of non-pharmacological treatment), or iii) an omission of an indicated medication.^35-37^ Improvement in the AOU was defined as a reduction of ≥1 prescribing omission. Several interrater reliability assessments showed moderate agreement between the outcome assessors when doing the MAI and AOU assessments.

#### Secondary outcomes

Secondary outcomes included medication appropriateness (as measured by the ordinal MAI), the number of prescribing omissions, long-term medications, falls and fractures, quality of life as measured by the EQ-5D-5L questionnaire,^38,39^ informal care received (e.g. unpaid care work by relatives/friends), and survival. For health economic analyses (reported separately) additional secondary outcomes were QALYs,^40^ health services use (e.g., number of GP and specialist visits), and direct medical costs accrued in one year.

## Data collection

Data was collected at baseline, 6 months, and 12 months. The data on medications, diagnoses, lab values, and vital data were imported from the EHR of patients through the FIRE database.^28^ Due to the substantial amount of missing data needed to assess the primary outcomes (∼35%), the study team collected missing information from participating GPs. The data on quality of life, health services use, falls and fractures were collected through phone calls with patients or legal representatives. GPs reported safety information on (serious) adverse events, including death.

### Sample size calculation

We calculated the sample size to test for superiority of the two primary outcome measures and used the Bonferroni approach to account for multiple testing. We assumed that 35% and 60% of patients will have an improvement in the MAI and that 10% and 30% will have an improvement in the AOU in the control and intervention group, respectively. Based on a two-sample comparison of proportions, a pre-specified number of GP clusters of 40 (20 per arm), and an ICC of 0.05, 7 patients were required per cluster to detect a difference in the proportion of improvement in the MAI score of 25% between the two groups with a power of 90% at a two-sided alpha-level of 0.025. Using the same assumption for the AOU, 7 patients per cluster were also required to detect a difference of 20%. This results in a total sample size of 280 patients (140 per arm). This sample size provides 81% power to detect a significant improvement in both the MAI score and the AOU index. To account for attrition due to dropout or death (15% estimated), we enlarged the number of patients per cluster to 8-10, with a final sample size of 320 patients total (160 per group).

### Statistical analysis

Sociodemographic characteristics of GPs were described. Patient characteristics were described and compared between groups using the Chi-squared test for categorical variables and the non-parametric Wilcoxon rank-sum test for continuous variables.^41^ The number of prescribing recommendations generated and implemented were descriptively analyzed.

In all model-based analyses (both intention-to-treat and per-protocol), multiple imputed data were used **(Appendix 2**). We used generalized estimating equation (GEE) models with robust standard errors to account for clustered data, which yields population-averaged effects.^42-44^ For binary outcomes, we computed odds ratios (OR) using a binomial distribution and a logit link. For count outcomes, we calculated incidence rate ratios (IRR) using a negative binomial distribution and a log link. For continuous outcomes, mean differences (MD) were computed using a Gaussian distribution and an identity link.

Several pre-specified subgroup comparisons were performed at GP and patient level through the models described above (**Appendix 3**), with additional interaction terms between randomization group and binary subgroup indicators. OR, IRR or MD for randomized comparisons are shown for each subgroup together with a p-value for interaction.

Due to differences in data exports from different EHR software programs, fewer medications and diagnoses were recorded for some patients. In secondary analyses, all models were re-run with the per-protocol set of patients. Here, we excluded all patients for whom <3 chronic conditions and <5 long-term medications were recorded, and all clusters that included <4 participants. In addition, we performed a post-hoc relaxed per-protocol analysis in which only patients with <1 chronic condition and <1 long-term medication recorded and clusters with <4 patients were excluded. In an additional sensitivity analysis, we adjusted models for potential confounders because cluster randomization may lead to imbalances in baseline characteristics between groups. Finally, we conducted an aggregated data analysis at the GP level.

All analyses were performed using Stata version 17.0^45^. We report the results in line with the Consolidated Standards of Reporting Trials extension for cluster trials.^46^ Deviations from the Statistical Analysis Plan are reported in **Appendix 4**.

### Patient and public involvement

GPs and patients aged ≥65 years with multimorbidity and polypharmacy were represented in the Safety and Data Monitoring Board (DSMB).

## Results

43 GPs working in Swiss primary care practices participated in the OPTICA trial (**eFigure 1**).^47^ Between January 2019 and February 2020, 323 older adults with multimorbidity and polypharmacy (median age: 77, interquartile range (IQR): 73-83, 45% women) gave their informed consent. 22 GPs with 160 patients were randomized to the intervention group, 21 GPs with 163 patients to the control group (**Figure 1**). During the 12-month follow-up period, 12 patients (3.7%) died, 12 patients (3.7%) were lost to follow-up and could not be reached for the data collection by phone, 5 patients (1.5%) opted out from the data collection by phone but agreed to be followed-up via the FIRE database, and 1 patient withdrew from the study.

**Figure 1.**
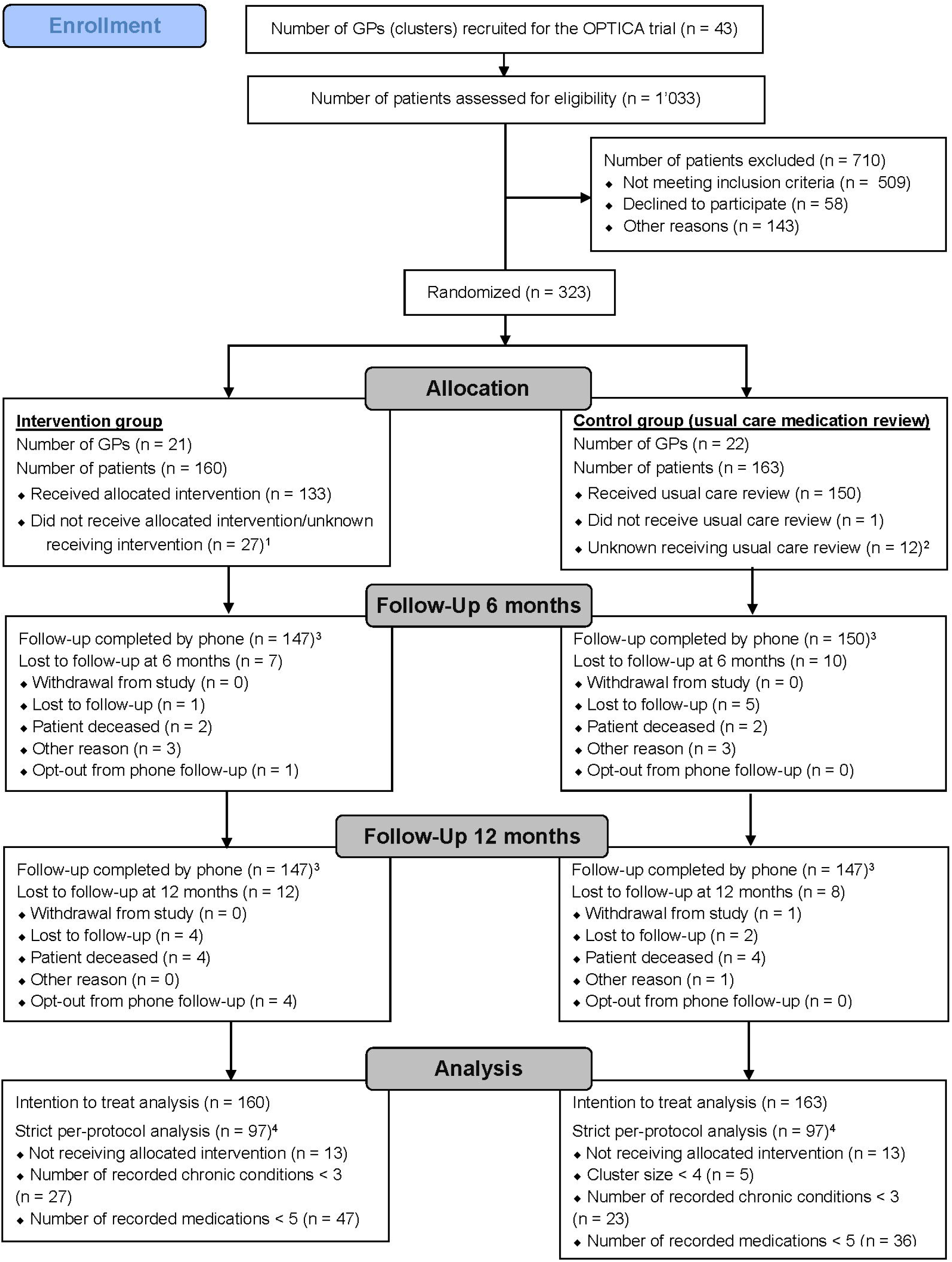
CONSORT patient flow chart. ^1^For these patients the drag/drop function in the ‘Systematic Tool to Reduce Inappropriate Prescribing’-Assistant (STRIPA) had not been used or was reset after the intervention. | ^2^ Reasons for this are that patients did not see their GP or had other urgent healthcare needs that had to be prioritized. | ^3^Referring to follow-up calls by phone. The time windows for these phone calls were: +15 days at baseline, ±30 days at the 6 months follow-up, and ±30 days at the 12 months follow-up. For all patients, except those who withdrew from the study, we could use the data from the FIRE database available (i.e., provided that the patient continued seeing the same GP). | ^4^Multiple criteria can apply. | Multiple imputed data were used for the analyses.

Participating GPs had a median age of 53 years (IQR: 44-58), median work experience of 14 years (IQR: 7-21), and 21% were female. At baseline, patients had a median number of long-term medications of 7 (IQR: 4-10) and a median number of chronic diagnoses of 7 (IQR: 4-10). The baseline characteristics of patients were similar in both groups, except for medication appropriateness, which was higher in the control group (**Table 1**).

**Table 1.** GP and patient characteristics at baseline.

| General practitioners (clusters) |  |  |  |  |
| --- | --- | --- | --- | --- |
|  | *All clusters*<br>*(n = 43)* | *Control group*<br>*(n = 22)* | *Intervention group*<br>*(n = 21)* |  |
|  | *median [lower and upper quartile] or n (%)* |  |  |  |
| Number of patients per cluster |  |  |  |  |
| <4 patients | 2 (5%) | 2 (9%) | 0 (0%) |  |
| 4-7 | 9 (21%) | 3 (14%) | 6 (29%) |  |
| ≥8 patients | 32 (74%) | 17 (77%) | 15 (71%) |  |
| Practice location |  |  |  |  |
| rural | 21 (49%) | 12 (55%) | 9 (43%) |  |
| urban/suburban | 22 (51%) | 10 (45%) | 12 (57%) |  |
| Age (in years) | 53 [44, 58] | 54 [44, 55] | 51 [44, 58] |  |
| Gender |  |  |  |  |
| male | 34 (79%) | 16 (73%) | 18 (86%) |  |
| female | 9 (21%) | 6 (27%) | 3 (14%) |  |
| Work experience as GP (in years) | 14 [7, 21] | 16 [7, 21] | 12 [8, 22] |  |
| Average number of consultations per workday | 25 [20, 28] | 25 [22, 30] | 25 [20, 25] |  |
| Practice form |  |  |  |  |
| individual practice | 7 (16%) | 5 (23%) | 2 (10%) |  |
| group practice | 36 (84%) | 17 (77%) | 19 (90%) |  |
| Average number of GPs in group practices | 3 [2, 4] | 3 [2, 4] | 4 [3, 5] |  |
| Patients |  |  |  |  |
|  | *All clusters*<br>*(n = 323)* | *Control*<br>*(n = 163)* | *Intervention*<br>*(n = 160)* | *p-value* |
| Age categories |  |  |  | 0.88 |
| 65-74 | 111 (34%) | 58 (36%) | 53 (33%) |  |
| 75-84 | 150 (46%) | 75 (46%) | 75 (47%) |  |
| ≥ 85 | 62 (19%) | 30 (18%) | 32 (20%) |  |
| Age (in years) | 77 [73, 83] | 77 [73, 83] | 77 [73, 83] | 0.76 |
| Gender |  |  |  | 0.82 |
| male | 177 (55%) | 88 (54%) | 89 (56%) |  |
| female | 146 (45%) | 75 (46%) | 71 (44%) |  |
| Highest education level |  |  |  | 0.57 |
| less than mandatory schooling | 4 (1.2%) | 1 (0.61%) | 3 (1.9%) |  |
| mandatory schooling | 118 (37%) | 62 (38%) | 56 (35%) |  |
| high school degree or apprenticeship | 145 (45%) | 70 (43%) | 75 (47%) |  |
| university or equivalent | 45 (14%) | 23 (14%) | 22 (14%) |  |
| other | 2 (0.62%) | 0 (0.00%) | 2 (1.3%) |  |
| Smoking status |  |  |  | 0.30 |
| smoker | 33 (10%) | 14 (8.6%) | 19 (12%) |  |
| past smoker | 140 (43%) | 65 (40%) | 75 (47%) |  |
| nonsmoker | 142 (44%) | 77 (47%) | 65 (41%) |  |
| Alcohol consumption in the 6 months prior to study enrolment |  |  |  | 0.91 |
| yes | 196 (61%) | 97 (60%) | 99 (62%) |  |
| Alcohol consumption in the 6 months prior to study enrolment (in units per week) | 2 [1, 7] | 2 [1, 7] | 2 [1, 7] | 0.87 |
| Permanent stay in nursing home |  |  |  | 0.29 |
| yes | 24 (7%) | 9 (6%) | 15 (9%) |  |
| Cognitive impairment |  |  |  | 0.77 |
| yes | 12 (4%) | 7 (4%) | 5 (3%) |  |
| Falls in the 6 months prior to study enrolment |  |  |  | 0.66 |
| yes | 58 (18%) | 27 (17%) | 31 (19%) |  |
| Number of falls in the 6 months prior to study enrolment | 1 [1, 1] | 1 [1, 1] | 1 [1, 1] | 0.54 |
| Number of hospitalizations in the 6 months prior to study enrolment | 0 [0, 0] | 0 [0, 0] | 0 [0, 0] | 0.46 |
| Number of long-term medications at baseline | 7 [4, 10] | 8 [5, 10] | 7 [4, 10] | 0.07 |
| Number of chronic conditions at baseline | 7 [4, 10] | 7 [4, 10] | 6 [4, 9] | 0.15 |
| Quality of life<sup>1</sup> | 75 [60, 80] | 70 [60, 80] | 75 [60, 80] | 0.61 |
| Pain or discomfort score (EQ-5D-5L) |  |  |  | 0.63 |
| no pain | 121 (37%) | 61 (37%) | 60 (38%) |  |
| slight pain | 64 (20%) | 31 (19%) | 33 (21%) |  |
| moderate pain | 84 (26%) | 41 (25%) | 43 (27%) |  |
| severe pain | 27 (8%) | 14 (9%) | 13 (8%) |  |
| extreme pain | 3 (1%) | 0 (0%) | 3 (2%) |  |
| Willingness to have medications deprescribed<sup>2</sup> |  |  |  | 0.72 |
| yes | 36 (11%) | 19 (12%) | 17 (11%) |  |
| Total Medication Appropriateness Index (MAI)<sup>3</sup> score at baseline | 12 [2, 38] | 5.0 [0, 38] | 15 [4, 38] | 0.024 |
| Averaged MAI score at baseline<sup>4</sup> | 1.9 [0.2, 5.2] | 0.6 [0.0, 4.9] | 3.0 [0.5, 5.4] | 0.03 |
| Total number of prescribing omissions<sup>5</sup> | 1 [0, 2] | 1 [0, 2] | 1 [0, 1] | 0.10 |
| Averaged number of prescribing omissions<sup>6</sup> | 0.1 [0.0, 0.2] | 0.1 [0.0, 0.2] | 0.1 [0.0, 0.2] | 0.33 |

*Missing values:* 0% for all GP characteristics, except for the number of consultations per day (2%). 0% for all patient characteristics except for 3% education level, 2% smoking status, 2% smoking consumption, 2% living situation, 4% falls, 3% number of hospitalizations, 8% for quality of life, 7% for both primary outcomes MAI and AOU at baseline. | <sup>1</sup>Measured by visual analogue scale of the European Quality of Life-5 Dimensions questionnaire (EQ-VAS). Values range from 0 to 100 with higher values indicating a higher quality of life. | <sup>2</sup>As measured by the question 'I would be willing to stop one or more of my medicines if my doctor said it was possible' of the 'revised Patients' Attitudes Towards Deprescribing' (rPATD) (Reeve et al. 2016). Dichotomized by agree/strongly agree vs don't know/disagree/strongly disagree. | <sup>3</sup>Adapted from Samsa, Hanlon, et al. *Journal of clinical epidemiology*. 1994. Sum of all MAI scores of each medication. Each individual MAI score ranges from 0 to 17 with higher values indicating a higher inappropriateness. | <sup>4</sup>Averaged by the number of chronic medications. | <sup>5</sup>As measured by the Assessment of Underutilization (AOU) based on Jeffery et al. *Consult Pharm*. 1999. Count of prescribing omissions for the chronic conditions at baseline. | <sup>6</sup>Averaged for the number of chronic conditions.

Per patient, an average of 5.4 prescribing recommendations were generated, thereof, an average of 3.7 were STOPP or START recommendations, (**Table 2**). On average, 1.0 START or STOPP recommendations were implemented per patient in the intervention group, with 58.5% of patients having had at least one recommendation implemented. The ten most common prescribing recommendations made by STRIPA are shown in **Table 3**.

**Table 2.**
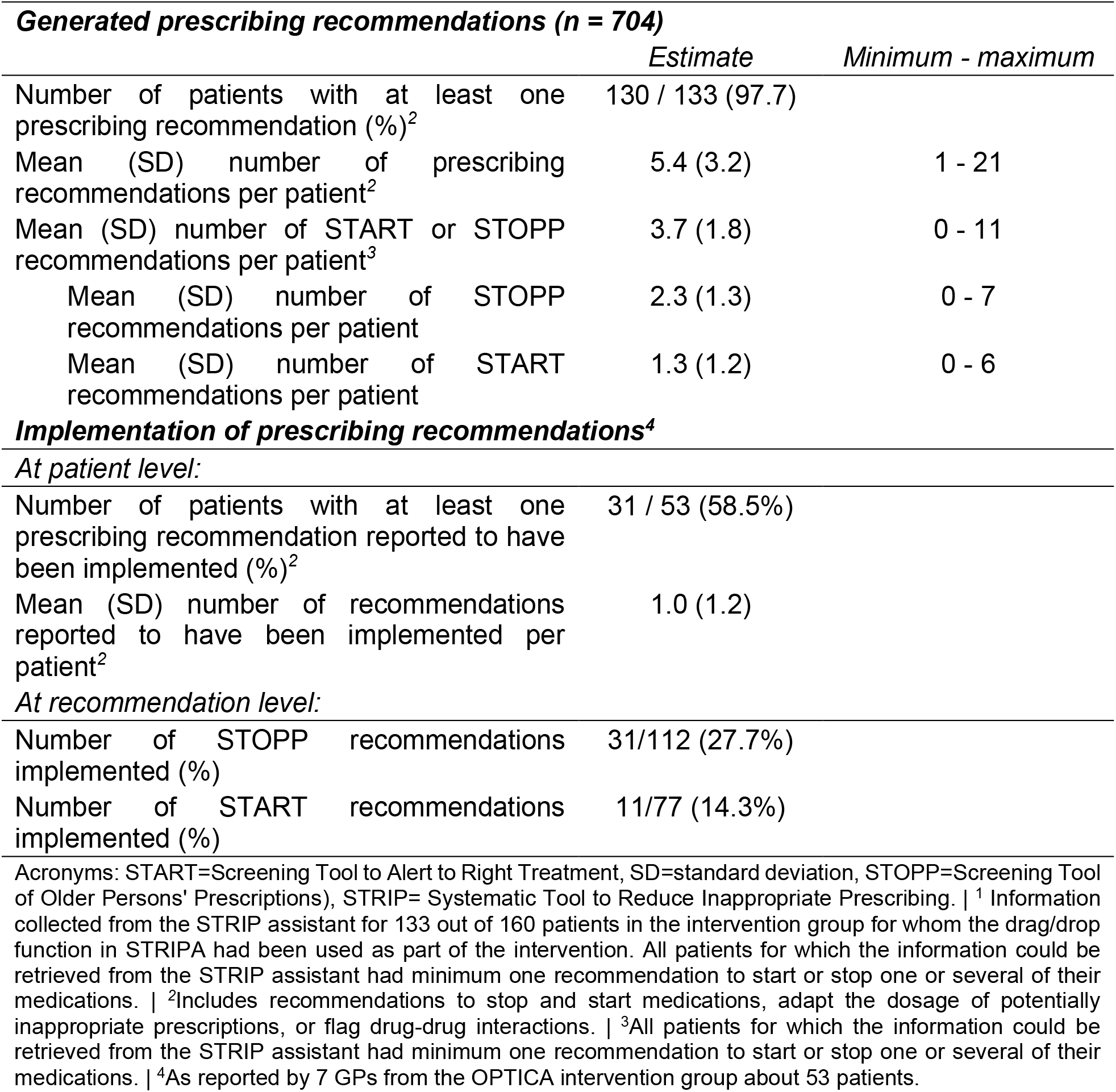
Recommendations generated by the ‘Systematic Tool to Reduce Inappropriate Prescribing’ to optimize prescribing in older adults (n = 133^1^)

|  | *Estimate* | *Minimum - maximum* |
| --- | --- | --- |
| Number of patients with at least one prescribing recommendation (%)<sup>2</sup> | 130 / 133 (97.7) |  |
| Mean (SD) number of prescribing recommendations per patient<sup>2</sup> | 5.4 (3.2) | 1 - 21 |
| Mean (SD) number of START or STOPP recommendations per patient<sup>3</sup> | 3.7 (1.8) | 0 - 11 |
| Mean (SD) number of STOPP recommendations per patient | 2.3 (1.3) | 0 - 7 |
| Mean (SD) number of START recommendations per patient | 1.3 (1.2) | 0 - 6 |

|  |  |
| --- | --- |
| Number of patients with at least one prescribing recommendation reported to have been implemented (%)<sup>2</sup> | 31 / 53 (58.5%) |
| Mean (SD) number of recommendations reported to have been implemented per patient<sup>2</sup> | 1.0 (1.2) |

***At recommendation level:***
|  |  |
| --- | --- |
| Number of STOPP recommendations implemented (%) | 31/112 (27.7%) |
| Number of START recommendations implemented (%) | 11/77 (14.3%) |

Acronyms: START=Screening Tool to Alert to Right Treatment, SD=standard deviation, STOPP=Screening Tool of Older Persons' Prescriptions), STRIP= Systematic Tool to Reduce Inappropriate Prescribing. | <sup>1</sup> Information
collected from the STRIP assistant for 133 out of 160 patients in the intervention group for whom the drag/drop function in STRIPA had been used as part of the intervention. All patients for which the information could be retrieved from the STRIP assistant had minimum one recommendation to start or stop one or several of their medications. | <sup>2</sup>Includes recommendations to stop and start medications, adapt the dosage of potentially inappropriate prescriptions, or flag drug-drug interactions. | <sup>3</sup>All patients for which the information could be retrieved from the STRIP assistant had minimum one recommendation to start or stop one or several of their medications. | <sup>4</sup>As reported by 7 GPs from the OPTICA intervention group about 53 patients.

**Table 3.**
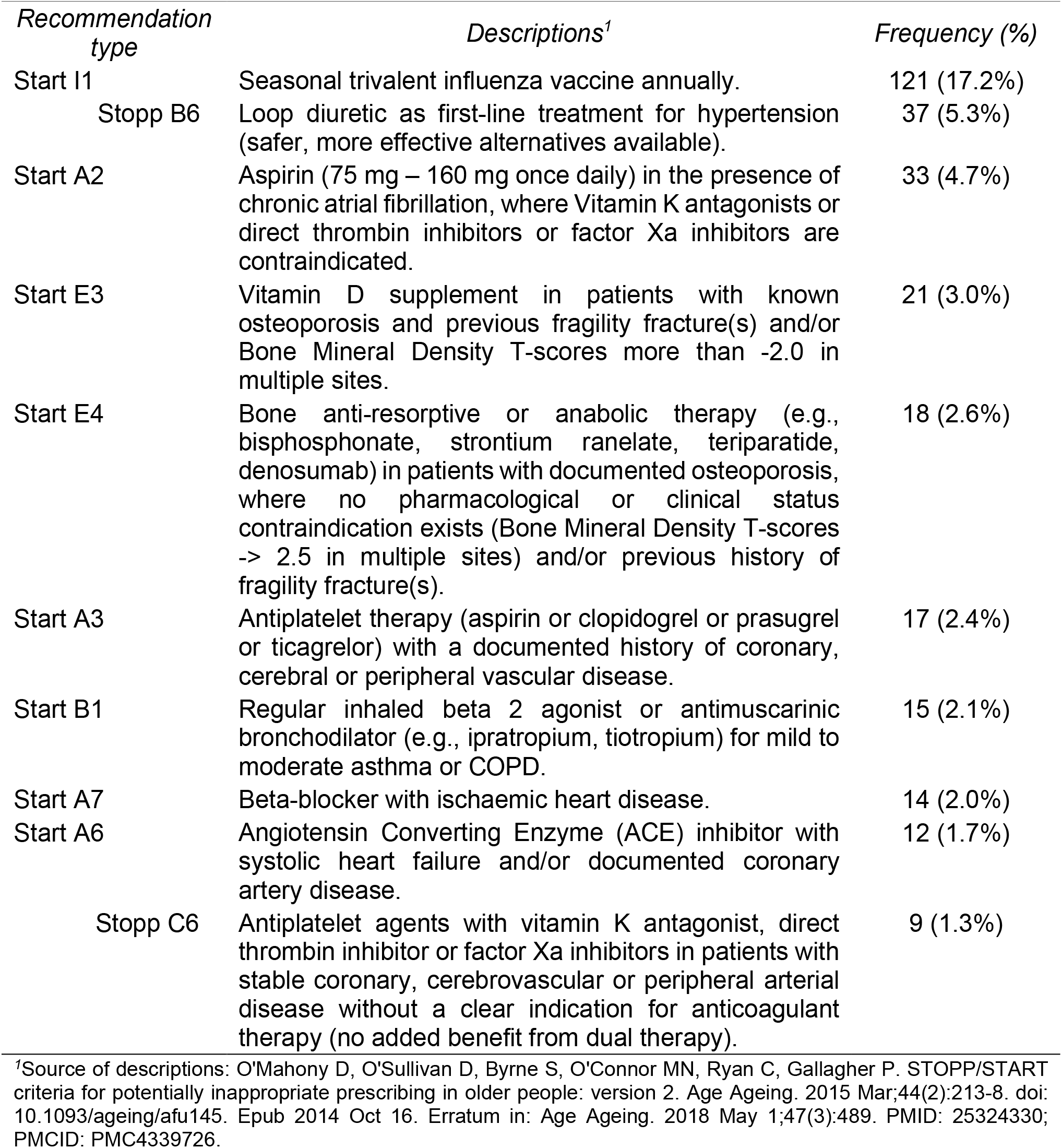
Ten most common prescribing recommendations generated by the ‘Systematic Tool to Reduce Inappropriate Prescribing’ Assistant of 704 recommendations made.

### Primary outcome measures

82% of patients had information on the Medication Appropriateness Index (MAI) for both the baseline and the 12-month follow-up and 87% of patients had information on prescribing omissions as assessed using the AOU for both timepoints (**eTable1**). 42% of patients had an improvement in the MAI between baseline and the 12-month follow-up and 14% had an improvement in the number of prescribing omissions (**eTable2**). In the supporting material, the development of the MAI score and the number of prescribing omissions are shown by timepoint (**eFigure3-5, eTable3**).

The analyses compared the intervention to the control group. In the intention-to-treat analysis, the odds ratio (OR) for an improvement in the MAI score (decrease by ≥1 score point) between baseline and the 12-month follow-up was 1.05 (95% confidence interval (CI): 0.59 to 1.87) and the OR for an improvement in the AOU (at least 1 prescribing omission less) was 0.90 (95% CI: 0.41 to 1.96) (**Table 4**). The sensitivity analysis adjusting for baseline characteristics did not show any significant group differences either (**eTable4**). Moreover, the intervention effect on medication appropriateness and prescribing omissions did not differ in subgroup analyses (**eFigure5**) nor in the aggregated data analysis (**eTable5**). The results based on the available case data did not show a difference either (**eTable6**).

**Table 4.** Outcomes: Comparison between the intervention and control group.

|  | Control<br>(n=163)<sup>4</sup> | Intervention<br>(n=160)<sup>4</sup> | Effect size | p-value |
| --- | --- | --- | --- | --- |
| **Primary outcomes** |  |  |  |  |
|  | *Number of events (%)* |  | *OR (95% CI)* |  |
| Improvement in the MAI score between baseline and 12-month follow-up | 67 (41%) | 68 (43%) | 1.05 (0.59 to 1.87) | 0.867 |
| Improvement in the number of prescribing omissions between baseline and 12-month follow-up | 28 (17%) | 24 (15%) | 0.90 (0.41 to 1.96) | 0.788 |
| **Secondary outcomes** |  |  |  |  |
| *Medication-related secondary outcomes* |  |  |  |  |
|  | *Number of events (%)* |  | *OR (95% CI)* |  |
| Improvement in the MAI score between baseline and 6-month follow-up | 64 (39%) | 68 (43%) | 1.14 (0.60 to 2.14) | 0.692 |
| Improvement in the number of prescribing omissions between baseline and 6-month follow-up | 28 (17%) | 19 (12%) | 0.67 (0.31 to 1.46) | 0.314 |
|  | *Mean (95% CI)* |  | *IRR (95% CI)<sup>5</sup>* |  |
| MAI total score at 6-month follow-up | 22 (17 to 27) | 25 (20 to 29) | 1.36 (0.89 to 2.08) | 0.153 |
| MAI total score at 12-month follow-up | 25 (19 to 30) | 26 (21 to 30) | 1.15 (0.74 to 1.79) | 0.528 |
| Total number of prescribing omissions at 6-month follow-up | 1.1 (0.9 to 1.3) | 1.1 (0.9 to 1.3) | 1.06 (0.83 to 1.36) | 0.641 |
| Total number of prescribing omissions at 12-month follow-up | 1.2 (1.0 to 1.3) | 0.9 (0.7 to 1.1) | 0.83 (0.65 to 1.07) | 0.151 |
|  | *Mean (95% CI)* |  | *MD (95% CI)<sup>5</sup>* |  |
| Number of medications at 6-month follow-up | 7.6 (7.0 to 8.2) | 7.2 (6.6 to 7.9) | 0.05 (-0.87 to 0.97) | 0.909 |
| Number of medications at 12-month follow-up | 8.0 (7.4 to 8.7) | 7.8 (7.2 to 8.4) | 0.26 (-0.64 to 1.16) | 0.576 |
| *Patient-reported secondary outcomes* |  |  |  |  |
|  | *Mean (95%-CI)* |  | *IRR (95% CI)* |  |
| Number of falls at 6-month follow-up | 0.3 (0.1 to 0.4) | 0.3 (0.2 to 0.4) | 0.96 (0.50 to 1.84) | 0.891 |

| Number of falls at 12-month follow-up | 0.2 (0.1 to 0.3) | 0.2 (0.1 to 0.3) | 0.90 (0.50 to 1.64) | 0.740 |
| --- | --- | --- | --- | --- |
|  | *n (%)* |  | *OR (95% CI)* |  |
| Any fracture(s) between baseline and 6-month follow-up<sup>1</sup> | 4 (3%) | 3 (2%) | 0.72 (0.17 to 3.10) | 0.660 |
| Any fracture(s) between baseline and 12-month follow-up<sup>1</sup> | 2 (1%) | 3 (2%) | 1.51 (0.27 to 8.50) | 0.643 |
| *Quality of life* | *Mean (95% CI)* |  | *MD (95% CI)<sup>5</sup>* |  |
| EQ-5D-5L utilities at 6-month follow-up<sup>2</sup> (inverse predicted utility – IPU) | 0.2 (0.2 to 0.2) | 0.2 (0.1 to 0.2) | -0.03 (-0.07 to 0.01) | 0.128 |
| EQ-5D-5L utilities at 12-month follow-up<sup>2</sup> (inverse predicted utility – IPU) | 0.1 (0.1 to 0.2) | 0.1 (0.1 to 0.2) | 0.00 (-0.04 to 0.03) | 0.933 |
| Visual analogue scale at 6-month follow-up<sup>3</sup> | 71 (68 to 74) | 72 (69 to 74) | 0.53 (-2.99 to 4.06) | 0.766 |
| Visual analogue scale at 12-month follow-up<sup>3</sup> | 73 (70 to 75) | 72 (70 to 75) | -0.42 (-3.77 to 2.93) | 0.807 |

Acronyms: AOU=Assessment of Underutilization, CI=Confidence Interval, IRR=Incident Rate Ratio, MAI=Medication Appropriateness Index, MD=Mean Difference, OR=Odds Ratio. This table is based on multiple imputed data. | <sup>1</sup> Due to the low number of fractures only the binary variable (yes/no) was considered. | <sup>2</sup> Calculated based on the German Value Set for the EQ-5D-5L by Ludwig et al. Pharmacoeconomics. 2018. | <sup>3</sup> Measured by visual analogue scale of the European Quality of Life-5 Dimensions questionnaire (EQ-VAS). Values range from 0 to 100 with higher values indicating a higher quality of life. | | <sup>4</sup> Number of patients with data (with multiple imputations). | <sup>5</sup> Adjusted for the respective baseline score. | Additional descriptive information on the secondary outcomes by study timepoint can be found in eTable11.

In the strict per-protocol analysis, the adjusted OR for an improvement in the MAI score between baseline and the 12-month follow-up was 1.03 (95% CI: 0.59 to 1.77) and the odds ratio for an improvement in the AOU was 1.25 (95% CI: 0.44 to 3.56) (**eTable7-8**). The relaxed per-protocol analysis showed similar results.

### Secondary outcomes

There was no difference between groups with regards to the secondary outcomes, when analyzing the primary outcomes – medication appropriateness and prescribing omissions – at the 6-month follow-up (**Table 4**), in the per-protocol analyses (**eTable9**) nor in additional medication-related outcomes (**eTable10**).

## Discussion

In this cluster randomized clinical trial evaluating the effect of a structured medication review intervention supported by an electronic decision support system in older adults with multimorbidity and polypharmacy, 58.5% of patients had at least one prescribing recommendation implemented. Despite the implementation of one STOPP or START recommendation per patient on average, there was no improvement in medication appropriateness nor a reduction in prescribing omissions after a 12-month follow-up period compared to usual care. While the intervention was not superior to the usual care, it could be safely delivered without causing any harm to patients.

Our null findings are in line with the literature on previous clinical trials testing the effect of medication review interventions on medication appropriateness in primary care settings. For instance, the PRIMUM trial, which randomized 72 primary care practices and 505 patients with multimorbidity and polypharmacy, did not find an improvement in the MAI after performing a medication review based on an eCDSS for GPs.^48^ This trial, however, did not assess under-prescribing and excluded patients with cognitive impairment. The results of the PRIMA-eDS trial with 359 primary care practices and 3,904 adults aged ≥75 years using multiple medications showed that the use of an eCDSS did not achieve a group difference in mortality or unplanned hospital admissions after a 24-month follow-up period.^25^ This study was strengthened by its longer observation period. However, it did not study medication underuse. In the OPTIMIZE trial, in which 3,012 patients with dementia or mild cognitive impairment from 19 primary care clinics were randomized, the number of (potentially inappropriate) medications were similar in both groups at the end of the 6-month follow-up period.^49^ These findings show that meaningful, sustainable improvements are difficult to achieve by the one-time use of eCDSS to support medication reviews in primary care settings.

Further, our findings are in line with the results of previous multicenter trials evaluating an eCDSS based on the STOPP/START criteria. For instance, the SENATOR trial, in which 1’536 inpatients were randomized, did not find any group difference concerning the recurrence of adverse drug events within 14 days of randomization.^26^ Similarly, in the OPERAM trial, in which 2’008 patients from 110 clusters were randomized and STRIPA was also used, the difference between the groups after a 12-month follow-up period was not statistically significant despite a trend towards a reduction of re-hospitalizations^19^. The intervention, however, came with potential cost-savings of CHF 3’588 per patient and a gain of 0.025 quality-adjusted life years (95% CI: -0.002 to 0.052) per patient.^50^ Despite the more user-friendly application of the STOPP/START criteria as an eCDSS, there are substantial issues related to implementing recommendations.

In the SENATOR trial, 15% of the overall recommendations were implemented.^26^ In the OPERAM trial, 62% of patients had ≥1 recommendation successfully implemented at 2 months.^19^ Our results from primary care settings are very similar to this. It would be unreasonable to expect every prescribing recommendation to be implementable, but the partial uptake of prescribing recommendations points to numerous issues that affect the overall effectiveness of medication review interventions. One reason for the partial uptake could be that not all the generated recommendations were relevant to the patients’ individual situations. For example, changes that had been tried in the past but did not work were recommended by the system. Poor integration of the decision-support system into the clinical workflow might have been another reason.^51^ Further, prescribing recommendations, particularly those related to stopping medications, have been shown to be difficult to implement due to numerous barriers faced by patients and prescribers.^52,53^

Future trials on medication optimization interventions would benefit from interventions focusing on overcoming implementation challenges. This not only requires preparatory qualitative studies to better understand the challenges, but also using implementation science approaches to better integrate interventions into clinical workflows, tailoring interventions to users’ needs, and piloting interventions. Further, future interventions may benefit from being designed as repeated interventions to accommodate the dynamic prescribing practices and frequent medication changes in older patients with multimorbidity and polypharmacy.

### Strengths and Limitations

The OPTICA trial has several strengths. First, it had minimal exclusion criteria, which resulted in the recruitment of patients that were comparable to other older patients with multimorbidity and polypharmacy in Swiss primary care.^47^ Second, patient recruitment was based on screening lists with random samples of each GPs’ patient population. Third, the randomization of GP clusters once the recruitment per cluster was completed, helped to limit differential recruitment. Fourth, a low number of patients were lost to follow-up due to the pragmatic data collection design consisting of phone calls and EHR data imports. The pragmatic nature of the trial in primary care settings provides real-world insights. Finally, the collection of information related to the implementation of prescribing recommendations provides additional insights. However, despite several reminders, we were unable to collect this information from all GPs.

We noted several limitations. First, due to the Hawthorne effect, GPs in the control group may have been more attentive to their prescribing practices and it cannot be rule out that some of them were diverging from their usual care. The number of medications, however, did not differ between the two groups at any timepoint. Second, the quality of the data exports from the FIRE database was challenging because there were differences in how different EHR software programs exported data. Therefore, the study team had to manually collect missing information from GP practices. Third, since the intervention occurred at a single time point, its effectiveness may have been diluted over time. Fourth, the implementation of an average of 1 STOPP/START recommendation per patient did not seem to affect the MAI. Next, the different barriers faced by patients and providers related to the implementation of prescribing recommendations, which are numerous and well-documented in the literature (e.g., fears of negative health outcomes, alert fatigue, etc.), may have led to a low implementation of recommendations. Finally, STRIPA used clinical information but did not consider patient preferences. This may also have contributed to patients’ reluctance to implement recommendations.

## Conclusion

In this primary care-based trial, 58.5% of patients had at least one prescribing recommendation implemented. Despite this, the medication review intervention centered around the use of an eCDSS did not lead to a significant improvement in medication appropriateness nor a reduction in prescribing omissions at 12 months compared to usual care. Nevertheless, the intervention could be safely delivered without causing any detriment to patient outcomes.

## Declarations

### Author Contributions

Dr. Katharina Jungo and Odile Stalder had full access to all the data in the study and take responsibility for the integrity of the data and the accuracy of the data analysis.

Concept and design: Prof. Nicolas Rodondi, Prof. Sven Streit, Prof. Matthias Schwenkglenks, PD Dr. Sven Trelle, Prof. Marco Spruit, Dr. Claudio Schneider, and Dr. Katharina Jungo designed the trial.

Acquisition, analysis, or interpretation of data: Dr. Katharina Jungo, Dr. Anna-Katharina Ansorg, Dr. Carmen Floriani, Dr. Zsofia Rozsnyai, Dr. Rahel Meier, Nathalie Schwab, Fabio Valeri, Dr. Michael Bagattini, PD Dr. Sven Trelle, Prof. Marco Spruit, Prof. Matthias Schwenkglenks, Prof. Nicolas Rodondi, and Prof. Sven Streit were involved in the data acquisition, analysis, and interpretation.

Drafting of the manuscript: Dr. Katharina Jungo and Odile Stalder wrote the first draft of the manuscript.

Critical revision of the manuscript for important intellectual content: All other authors provided feedback and approved the final version of the manuscript.

Statistical analysis: Odile Stalder, Dr. Andreas Limacher, and PD Dr. Sven Trelle provided statistical support and conducted the statistical analyses.

Obtained funding: Prof. Nicolas Rodondi, Prof. Matthias Schwenkglenks and Prof. Sven Streit obtained the funding for the OPTICA trial.

Administrative, technical, or material support: Nathalie Schwab and Fabio Valeri provided administrative and technical support.

Supervision: Prof. Nicolas Rodondi and Prof. Sven Streit supervised the conduct of the trial.

## Supporting information

CONSORT Checklist

## Data Availability

We will make the data for this study available to other researchers upon request after publication. The data will be made available for scientific research purposes, after the proposed analysis plan has been approved. Data and documentation will be made available through a secure file exchange platform after approval of the proposal. In addition, a data transfer agreement has to be signed (which defines obligations that the data requester must adhere to with regard to privacy and data handling). Deidentified participant data limited to the data used for the proposed project will be made available, along with a data dictionary and annotated case report forms. For data access, please contact the corresponding author.

## Funding/Support

The OPTICA trial was supported by the Swiss National Science Foundation, within the framework of the National Research Programme 74 “Smarter Health Care” (NRP74) under contract number 407440_167465 (to Prof. Streit, Prof. Rodondi, and Prof. Schwenkglenks).

## Acknowledgements

We would like to thank the GPs who participated in the OPTICA trial for their great contributions without which it would have been impossible to conduct this study. We also thank the participants for consenting to participate in our study. We would like to thank Shana Bertato, Rebekah Hoeks, Dominic Salvisberg, Yasmin Mohd Yatim, Melisa Harmanci, und Linda Chau for their support with the data entry. We would like to thank Dr. Kristie Weir for her editorial suggestions. Katharina Tabea Jungo is a member of the Junior Investigator Intensive Program of the US Deprescribing Research Network, which is funded by the National Institute on Aging.

## Ethical approval

The study protocol of the OPTICA trial and other documentation was approved by the competent ethics committee of the canton of Bern (KEK), Switzerland, and the Swiss regulatory authority (Swissmedic) (BASEC ID: 2018–00914). The KEK and Swissmedic received annually safety reports and were informed about the end of the study. All participants gave their written informed consent. The OPTICA trial was performed in accordance with relevant regulations and guidelines.

