## Supporting Material for "Optimizing Prescribing in Older Adults with Multimorbidity and Polypharmacy in Primary Care: A Cluster Randomized Clinical Trial (OPTICA Trial)"

### Appendix 1. Details on the assessment of the Medication Appropriateness Index (MAI).

We assessed the AOU for each non-acute condition of the patients and the MAI for each long-term medication (≥90 days, no stop date; “as needed” medications were excluded). In this trial, we used the 10-item version of the MAI, however, we excluded the cost-effectiveness item for feasibility reasons. Using data on medications, diagnoses, and lab values the assessors rated the nine remaining criteria of the MAI for each medication using a three-point scale ranging from A=appropriate, B=marginally appropriate, to C=inappropriate. Each of the nine criteria has a weight of 1-3.[1] Each “inappropriate” rating received the respective weight, while each “marginally appropriate” and “appropriate” rating was weighted with 0. This resulted in a score from 0 to 17 for each medication.

### Appendix 2. Multiple imputation.

We used multiple imputation by chained equations to impute missing values in primary outcomes (MAI score and AOU index) at baseline, 6 and 12 months. Imputation models were based on all baseline characteristics of the patients and some GPs characteristics (**see below**). All these imputed variables were used to impute all the secondary outcomes at 6 and 12 months. Predictive mean matching (pmm) and logit models were used to impute ordinal and binary variables, respectively. In total, fifty imputed data sets were generated, which were analyzed as described above using Rubin’s rules to combine results across data sets.[2]

*Variables included in the multiple imputation model:*

**GP-level variables:** randomization group, practice form, practice size, canton of the GP practice, GPs’ work experience in years, and practice location

**Patient-level variables:** age, sex, education level, smoking status, alcohol consumption, permanent nursing home stay, number of falls, number of hospitalizations, number of chronic medications, number of chronic conditions, quality of life predicted utilities at baseline, EQ-5D visual analogue scale, and patients’ willingness to have medications deprescribed.

### Appendix 3. Deviations from the Statistical Analysis Plan (SAP)

Rather than using a random effects Poisson model for secondary count outcomes, we used a robust GEE model with a negative binomial distribution and a log link. The Poisson distribution is a special case of the negative binomial distribution and is too restrictive in this case, due to overdispersion. Furthermore, the GEE approach is more consistent with the other analyses than the random effects model.

Secondary continuous outcomes (MAI score, AOU index) were considered as count data and were analyzed using a GEE model with a negative binomial distribution and a log link due to their skewed distribution. Since the MAI score only consists of integers with a high proportion of zeros, its distribution cannot be considered as Gaussian.

We did not run the subgroup analysis related to medication adherence, since the data on medication adherence had not been collected at baseline but at the 12-month follow-up.

The rest of the analysis is consistent with the principal features of the statistical methods described in the statistical analysis plan (SAP).

### Appendix 4. Deviations from the Statistical Analysis Plan (SAP)

At the GP level, the following subgroup analyses were performed: age in years (<median, ≥median), experience in years (<median, ≥median), practice size (single vs. group practice), and sex (male vs. female).

At the patient level, we performed the following subgroup analyses: number of medications (<10, ≥10), sex (male vs. female), living conditions (nursing home vs. community dwelling), age in years (65-74, 75-84, ≥85), number of chronic conditions (<median, ≥median), and willingness to deprescribe (question from rPATD ‘If my doctor said it was possible, I would be willing to stop one or more of my regular medicines’ – dichotomized by strongly agree/agree vs. unsure, disagree/strongly disagree).

### eFigure 1. Screening and recruitment flow chart of general practitioners and patients in the OPTICA trial, a cluster-randomized controlled trial in Swiss primary care settings.

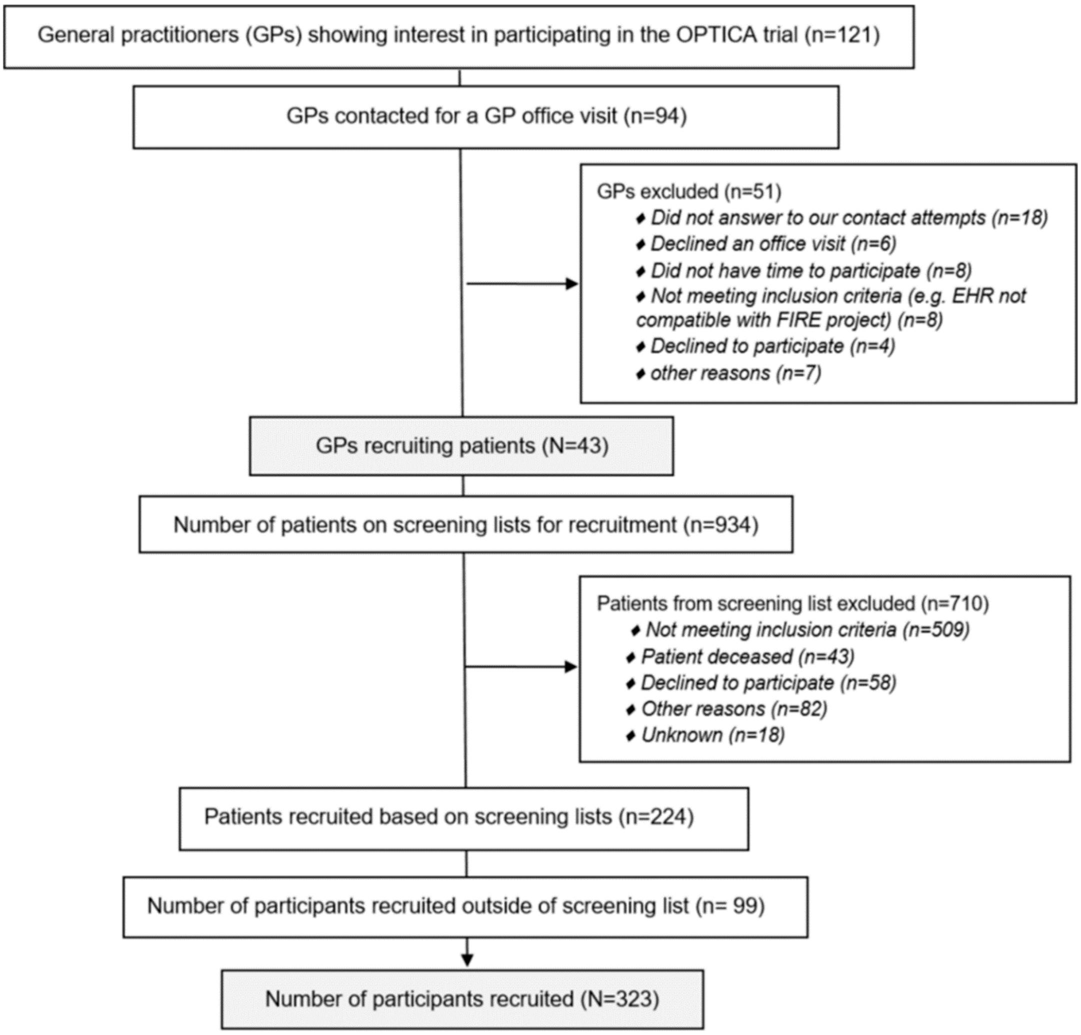

Abbreviation: OPTICA = Optimizing PharmacoTherapy in older multimorbid adults In primary CAre. | Source: Jungo KT, Meier R, Valeri F, Schwab N, Schneider C, Reeve E, Spruit M, Schwenkglenks M, Rodondi N, Streit S. Baseline characteristics and comparability of older multimorbid patients with polypharmacy and general practitioners participating in a randomized controlled primary care trial. BMC Fam Pract. 2021 Jun 22;22(1):123. doi: 10.1186/s12875-021-01488-8. PMID: 34157981; PMCID: PMC8220761.

### eTable 1. Availability of the information on the Medication Appropriateness Index (MAI) and the Assessment of Underutilization (AOU)

| | All participants (n = 323) | Control group (n=163) | Intervention group (n=160) |
|---|---|---|---|
| **AOU availability**[1] | | | |
| Baseline and 12-month follow-up | 265 (82%) | 133 (82%) | 132 (82%) |
| Only baseline | 27 (8.4%) | 14 (8.6%) | 13 (8.1%) |
| Only 12-month follow-up | 6 (1.9%) | 5 (3.1%) | 1 (0.6%) |
| Baseline and 12-month follow-up not available | 25 (7.7%) | 11 (6.7%) | 14 (8.7%) |
| **MAI availability**[2] | | | |
| Baseline and 12-month follow-up | 282 (87%) | 143 (88%) | 139 (87%) |

| | | | |
|---|---|---|---|
| Only baseline | 19 (5.9%) | 9 (5.5%) | 10 (6.3%) |
| Only 12-month follow-up | 4 (1.2%) | 3 (1.8%) | 1 (0.6%) |
| Only 6-month follow-up available | 14 (4.3%) | 5 (3.1%) | 9 (5.6%) |
| Baseline and 12-month follow-up not available | 4 (1.2%) | 3 (1.8%) | 1 (0.6%) |

This table is based on the raw data. | [1]The AOU was assessed for each chronic condition at the different timepoints. Based on Jeffery et al. Consult Pharm. 1999. | [2]The MAI was evaluated for each chronic medication at the different timepoints. Adapted from Samsa, Hanlon, et al. Journal of clinical epidemiology. 1994.

### eTable 2. Improvement of the primary outcomes: Medication Appropriateness Index (MAI) and Assessment of Underutilization (AOU)

| | **All participants (n = 323)** | **Control group (n=163)** | **Intervention group (n=160)** |
|---|---|---|---|
| **Improvement in MAI** | | | |
| No | 163 (50%) | 86 (53%) | 77 (48%) |
| Yes | 119 (37%) | 57 (35%) | 62 (39%) |
| Missing | 41 (13%) | 20 (12%) | 21 (13%) |
| **Improvement in AOU** | | | |
| No | 227 (70%) | 111 (68%) | 116 (73%) |
| Yes | 38 (12%) | 22 (13%) | 16 (10%) |
| Missing | 58 (18%) | 30 (18%) | 28 (18%) |

This table is based on the raw data.

### eTable 3. Descriptive primary outcomes.

| | **Control group (n = 163)** | | **Intervention group (n = 160)** | |
|---|---|---|---|---|
| | Missing n (%) | median [LQ, UQ]; mean (SD) or n (%) | Missing n (%) | median [LQ, UQ]; mean (SD) or n (%) |
| *Medication Appropriateness Index* | | | | |
| MAI score total (baseline) | 11 (7%) | 5.0 [0.00, 38]; 27 (43) | 11 (7%) | 15 [4.0, 38]; 26 (28) |
| MAI score total (6-month follow-up) | 20 (12%) | 7.0 [0.00, 36]; 22 (31) | 20 (13%) | 14 [3.0, 38]; 25 (27) |
| MAI score total (12-month follow-up) | 14 (9%) | 8.0 [0.00, 36]; 24 (33) | 19 (12%) | 15 [3.0, 38]; 26 (26) |
| MAI improvement between baseline and 12-month follow-up | 20 (12%) | | 21 (13%) | |
| no | | 86 (53%) | | 77 (48%) |
| yes | | 57 (35%) | | 62 (39%) |
| *Assessment of underutilization* | | | | |
| Number of prescribing omissions (baseline) | 16 (10%) | 1.0 [0.0, 2.0]; 1.2 (1.2) | 15 (9%) | 1.0 [0.0, 1.0]; 0.99 (1.3) |
| Number of prescribing omissions (6-month follow-up) | 30 (18%) | 1.0 [0.0, 2.0]; 1.1 (1.2) | 25 (16%) | 1.0 [0.0, 2.0]; 1.0 (1.2) |
| Number of prescribing omissions (12-month follow-up) | 25 (15%) | 1.0 [0.0, 2.0]; 1.2 (1.1) | 27 (17%) | 1.0 [0.0, 1.0]; 0.87 (1.1) |
| Improvement in the AOU between baseline and 12-month follow-up | 30 (18%) | | 28 (18%) | |
| no | | 111 (68%) | | 116 (73%) |
| yes | | 22 (13%) | | 16 (10%) |

Acronyms: LQ=lower quartile, UQ=upper quartile, SD=standard deviation, MAI=Medication Appropriateness Index, AOU=Assessment of underutilization. | This table is based on the raw data.

### eFigure 2. Development of medication appropriateness over time: Means and 95%-confidence intervals for the total Medication Appropriateness Index Score (MAI) score by study timepoint

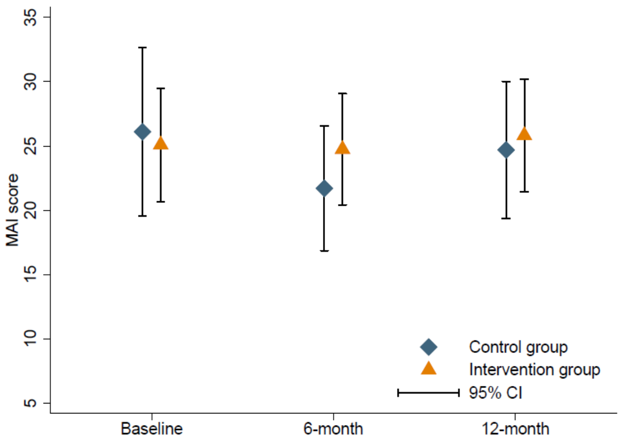

The numbers in this figure were calculated using multiple imputed data.

### eFigure 3. Development of prescribing omissions over time: Means and 95%-confidence intervals for the total number of prescribing omissions as measured by the Assessment of Underutilization (AOU) by study timepoint

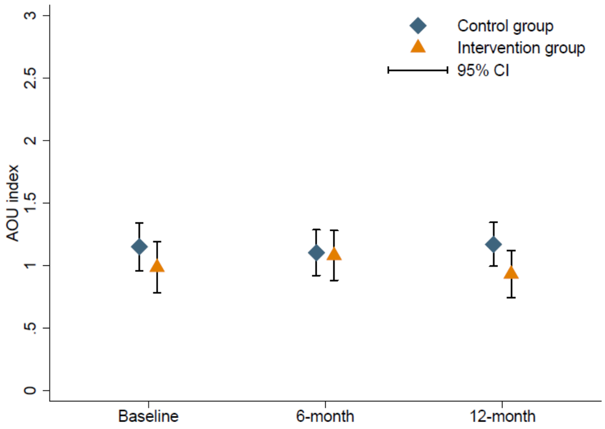

The numbers in this figure were calculated using multiple imputed data.

**eFigure 4. Primary outcomes by timepoint.**

**Medication Appropriateness Index Score (MAI)**

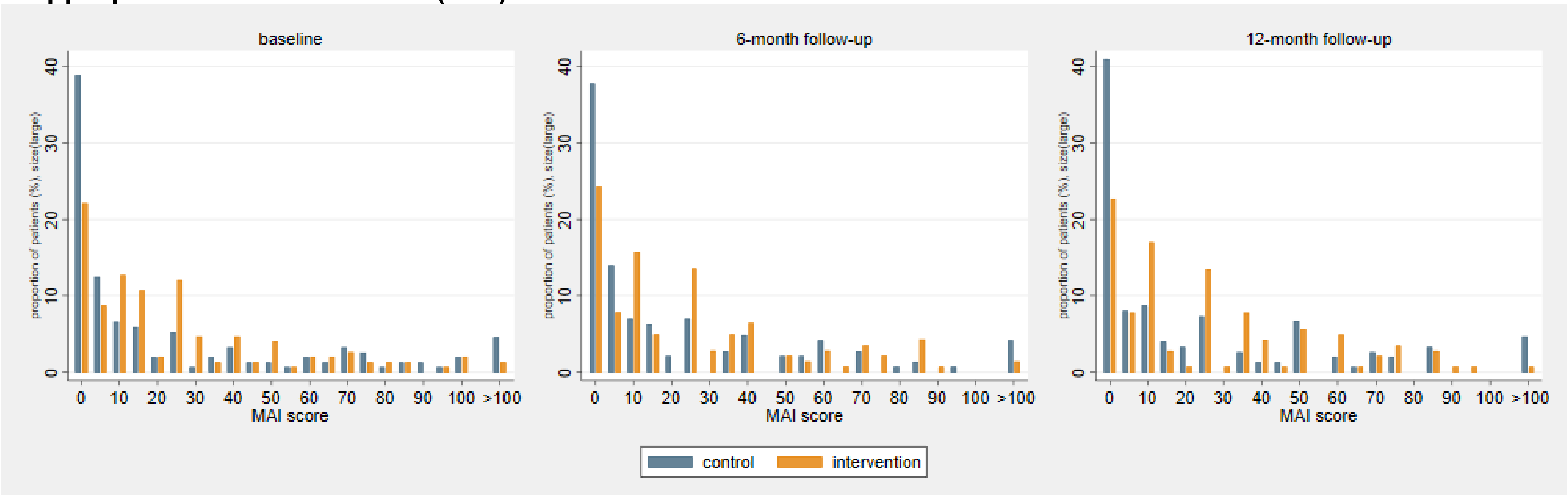

**Assessment of Underutilization (AOU)**

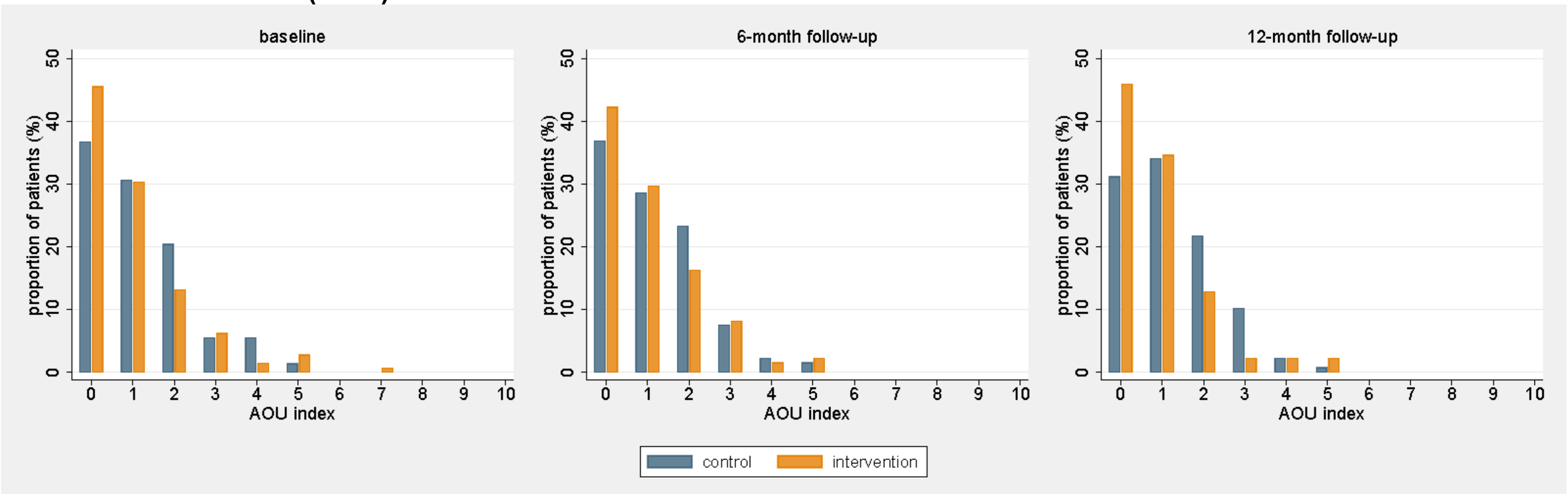

### eTable 4. Adjusted primary outcome models

| *4a. Adjusted for baseline characteristics* | | | | |
|---|---|---|---|---|
| | *Control (n=163)* | *Intervention (n=160)* | *OR (95% CI)* | *p-value* |
| | *n (%)* | | | |
| Improvement in the MAI score between baseline and 12-month follow-up | 57 (40%) | 62 (45%) | 1.01 (0.58 to 1.75) | 0.971 |
| Improvement in the AOU between baseline and 12-month follow-up | 22 (17%) | 16 (12%) | 1.07 (0.43 to 2.64) | 0.881 |
| ***4b. Adjusted for all subgroup variables[1]*** | | | | |
| | *n (%)* | | *OR (95% CI)* | |
| Improvement in the MAI score between baseline and 12-month follow-up | 67 (41%) | 68 (43%) | 1.36 (0.73 to 2.53) | 0.326 |
| Improvement in the AOU between baseline and 12-month follow-up | 28 (17%) | 24 (15%) | 0.91 (0.35 to 2.39) | 0.845 |

AOU=Assessment of Underutilization, CI=Confidence Interval, MAI=Medication Appropriateness Index, OR=Odds Ratio. | Calculated using multiple imputed data. | [1]Models were adjusted for number of chronic medications, patient's gender, permanent nursing home stay (yes/no), number of chronic conditions, patients' willingness to have medications deprescribed, patients' age, practice form, GPs' gender, GPs' age, and GPs' experience.

### eFigure 5. Subgroup analyses for the primary outcomes

(n = 323)

*Part A) Improvement in medication appropriateness between baseline and 12-month follow-up as measured by the Medication Appropriateness Index (MAI)*

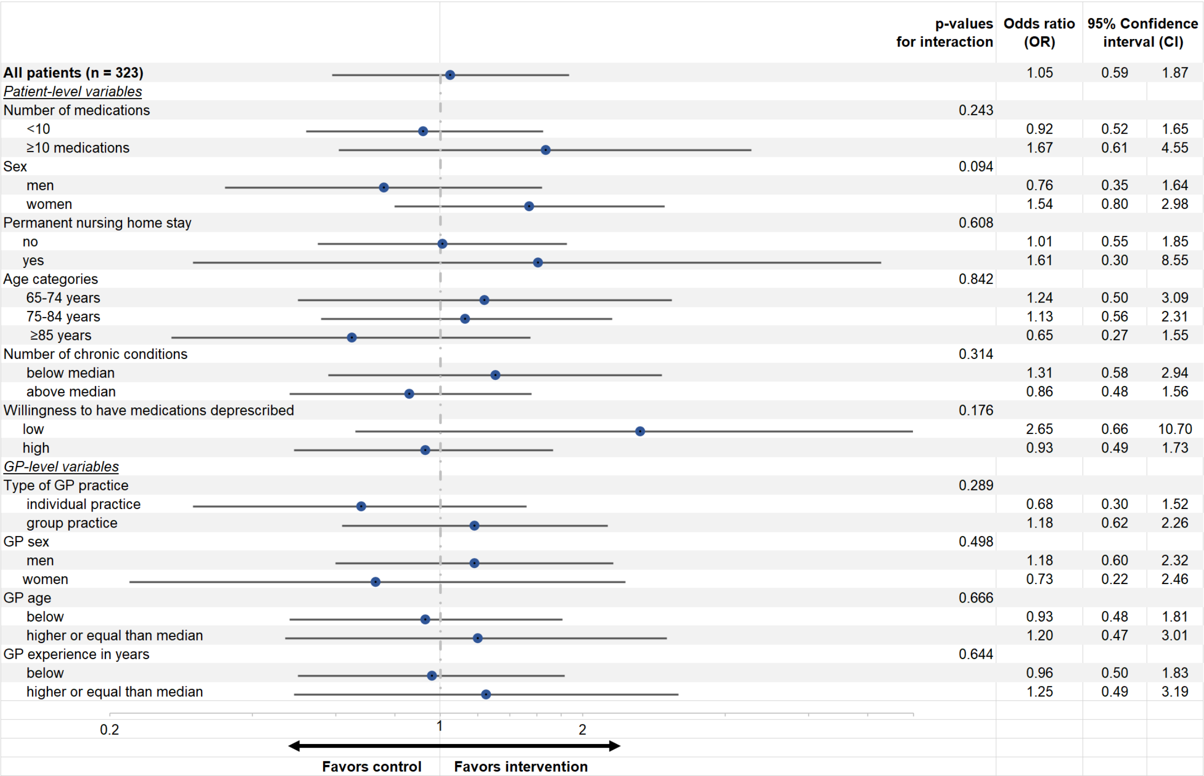

*Part B) Improvement in prescribing omissions between baseline and 12-month follow-up as measured by the Assessment of Underutilization*

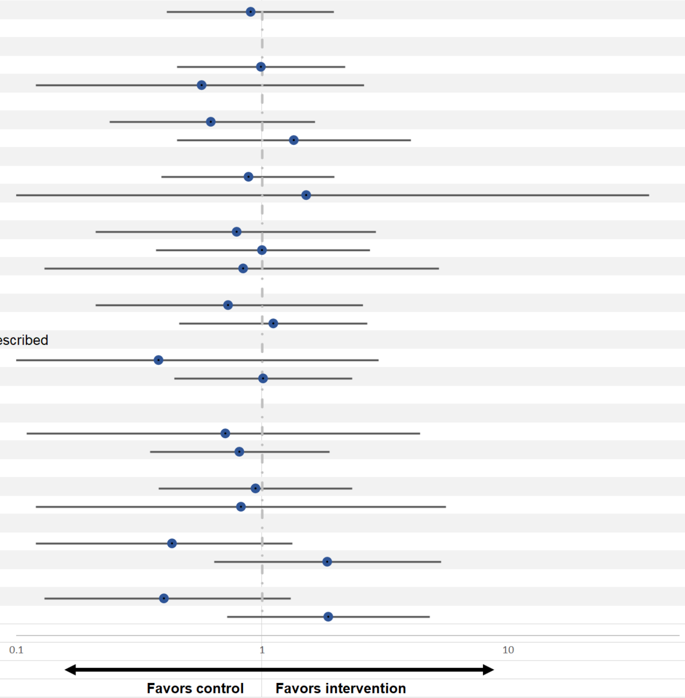

| | p-values for interaction | Odds ratio (OR) | 95% Confidence interval (CI) | |
|---|---|---|---|---|
| **All patients (n = 323)** | | 0.90 | 0.41 | 1.96 |
| *Patient-level variables* | | | | |
| Number of medications | 0.447 | | | |
| <10 | | 0.99 | 0.45 | 2.18 |
| ≥10 medications | | 0.57 | 0.12 | 2.60 |
| Sex | 0.234 | | | |
| men | | 0.62 | 0.24 | 1.64 |
| women | | 1.35 | 0.45 | 4.02 |
| Permanent nursing home stay | 0.743 | | | |
| no | | 0.88 | 0.39 | 1.97 |
| yes | | 1.51 | 0.06 | 37.44 |
| Age categories | 0.737 | | | |
| 65-74 years | | 0.79 | 0.21 | 2.90 |
| 75-84 years | | 1.00 | 0.37 | 2.74 |
| ≥85 years | | 0.84 | 0.13 | 5.24 |
| Number of chronic conditions | 0.542 | | | |
| below median | | 0.73 | 0.21 | 2.57 |
| above median | | 1.11 | 0.46 | 2.68 |
| Willingness to have medications deprescribed | 0.380 | | | |
| low | | 0.38 | 0.05 | 2.97 |
| high | | 1.01 | 0.44 | 2.32 |
| *GP-level variables* | | | | |
| Tpye of GP practice | 0.891 | | | |
| individual practice | | 0.71 | 0.11 | 4.39 |
| group practice | | 0.81 | 0.35 | 1.88 |
| GP sex | 0.897 | | | |
| men | | 0.94 | 0.38 | 2.32 |
| women | | 0.82 | 0.12 | 5.60 |
| GP age | 0.067 | | | |
| below | | 0.43 | 0.12 | 1.33 |
| higher or equal than median | | 1.84 | 0.64 | 5.34 |
| GP experience in years | 0.047 | | | |
| below | | 0.40 | 0.13 | 1.31 |
| higher or equal than median | | 1.86 | 0.72 | 4.80 |

Sub-group analyses as specified in the Statistical Analysis Plan. | Forest plots displayed on a logarithmic scale.

### eTable 5. Aggregated data analysis.

| | *Control (n=22 GPs)* | *Intervention (n=21 GPs)* | *Percentage mean difference (95% CI)* | *p-value* |
|---|---|---|---|---|
| | *Mean percentage (95%-CI)* | | | |
| Improvement in the MAI score between baseline and 12-month follow-up | 41.0 (26.9 to 55.2) | 41.6 (30.8 to 52.5) | 0.61 (-16.76 to 17.98) | 0.944 |
| Improvement in the AOU between baseline and 12-month follow-up | 19.0 (7.2 to 30.8) | 13.4 (6.1 to 20.7) | -5.64 (-19.45 to 8.18) | 0.415 |

AOU=Assessment of Underutilization, CI=Confidence Interval, MAI=Medication Appropriateness Index, OR=Odds Ratio. | Calculated using multiple imputed data.

### eTable 6. Available case analysis: Primary and secondary outcomes. (n = 271)

| | *Control* | *Intervention* | | *p-value* |
|---|---|---|---|---|
| **Primary outcomes** | *n (%)* | | *OR (95% CI)* | |
| Improvement in the MAI score between baseline and 12-month follow-up | 57/143 (40%) | 62/139 (45%) | 1.19 (0.67 to 2.12) | 0.552 |
| Improvement in the AOU between baseline and 12-month follow-up | 22/133 (17%) | 16/132 (12%) | 0.72 (0.32 to 1.63) | 0.430 |

| **Secondary outcomes** | | | | |
|---|---|---|---|---|
| *Medication-related secondary outcomes* | | | | |
| | *n (%)* | | *OR (95% CI)* | |
| Improvement in the MAI score between baseline and 6-month follow-up | 53/140 (38%) | 62/139 (45%) | 1.34 (0.69 to 2.63) | 0.389 |
| Improvement in the AOU between baseline and 6-month follow-up | 19/128 (15%) | 15/134 (11%) | 0.74 (0.32 to 1.70) | 0.479 |
| | *Mean (95% CI)* | | *IRR (95% CI)* [4] | |
| MAI total score at 6-month follow-up | 21.8 (16.7; 27.0) | 25.3 (20.9; 29.8) | 1.33 (0.85 to 2.07) | 0.216 |
| MAI total score at 12-month follow-up | 23.6 (18.2; 29.0) | 26.3 (21.9; 30.7) | 1.11 (0.68 to 1.81) | 0.682 |
| Total number of prescribing omissions at 6-month follow-up as measured by the AOU | 1.1 (0.9; 1.3) | 1.0 (0.8; 1.2) | 1.02 (0.78 to 1.33) | 0.878 |
| Total number of prescribing omissions at 12-month follow-up as measured by the AOU | 1.2 (1.0; 1.4) | 0.9 (0.7; 1.1) | 0.82 (0.64 to 1.06) | 0.136 |
| | *Mean (95% CI)* | | *MD (95% CI)*[4] | |
| Number of medications at 6-month follow-up | 8 (7; 8) | 7 (7; 8) | 0.32 (-0.33 to 0.96) | 0.334 |
| Number of medications at 12-month follow-up | 8 (7; 8) | 8 (7; 9) | 0.43 (-0.38 to 1.23) | 0.300 |
| *Patient-reported secondary outcomes* | | | | |
| | *Mean (95% CI)* | | *IRR (95% CI)*[4] | |
| Number of falls at 6-month follow-up | 0.3 (0.1; 0.4) | 0.3 (0.2; 0.4) | 0.92 (0.46 to 1.83) | 0.816 |
| Number of falls at 12-month follow-up | 0.2 (0.1; 0.3) | 0.2 (0.1; 0.3) | 0.84 (0.47 to 1.51) | 0.566 |
| | n (%) | | *OR (95% CI)* | |
| Any fracture(s) between baseline and 6-month follow-up[1] | 4 (3%) | 3 (2%) | 0.72 (0.17 to 3.10) | 0.660 |
| Any fracture(s) between baseline and 12-month follow-up[1] | 2 (1%) | 3 (2%) | 1.51 (0.27 to 8.50) | 0.643 |
| *Quality of life* | | | | |
| | *Mean (95% CI)* | | *MD (95% CI)* [4] | |
| EQ-5D-5L utilities at 6-month follow-up[2] (inverse predicted utility – IPU) | 0 (0 to 0) | 0 (0 to 0) | -0.03 (-0.07 to 0.01) | 0.139 |
| EQ-5D-5L utilities at 12-month follow-up[2] (inverse predicted utility – IPU) | 0 (0 to 0) | 0 (0 to 0) | 0.00 (-0.03 to 0.04) | 0.915 |
| Visual analogue scale at 6-month follow-up[3] | 71 (68; 74) | 72 (69; 75) | 0.88 (-2.43 to 4.18) | 0.603 |
| Visual analogue scale at 12-month follow-up[3] | 73 (70; 75) | 73 (71; 76) | -0.09 (-3.28 to 3.09) | 0.955 |

Acronyms: AOU=Assessment of Underutilization, CI=Confidence Interval, IRR=Incident Rate Ratio, MAI=Medication Appropriateness Index, MD=Mean Difference, OR=Odds Ratio. This table is based on multiple imputed data. | [1]Due to the low number of fractures only the binary variable (yes/no) was considered. | [2]Calculated based on the German Value Set for the EQ-5D-5L by Ludwig et al. PharmacoEconomics. 2018. | [3]Measured by visual analogue scale of the European Quality of Life-5 Dimensions questionnaire (EQ-VAS). Values range from 0 to 100 with higher values indicating a higher quality of life. | [4]Adjusted for baseline characteristics.

### eTable 7. Per-protocol set of participants.

***Strict per-protocol set of patients.*** [1]

| | *Randomized (n = 323)* | *Intervention (n = 160)* | *Control (n = 163)* |
|---|---|---|---|
| Not receiving allocated intervention | 26 | 13 | 13 |
| Cluster <4 | 5 | 0 | 5 |
| Age <65 years | 0 | 0 | 0 |
| Number of recorded chronic conditions <3 | 50 | 27 | 23 |
| Number of recorded medications <5 | 83 | 47 | 36 |
| PPset total (n = 194) | 194 | 97 | 97 |

***Relaxed per-protocol set of patients.*** [1]

| | *Randomized (n = 323)* | *Intervention (n = 160)* | *Control (n = 163)* |
|---|---|---|---|
| Not receiving intervention | 26 | 13 | 13 |
| Cluster <4 | 5 | 0 | 5 |
| Age <65 years | 0 | 0 | 0 |
| Number of recorded chronic conditions <1 | 31 | 15 | 16 |
| Number of recorded medications <1 | 22 | 11 | 11 |
| PPset total (n = 261) | 261 | 138 | 123 |

[1] Multiple criteria can apply.

### eTable 8. Per-protocol analyses: Primary outcomes

| | *Control* | *Intervention* | *OR (95% CI)* | *p-value* |
|---|---|---|---|---|
| **Strict per-protocol analysis** *(n = 194, clusters = 39)* | *N=97* | *N= 97* | | |
| Improvement in the MAI score between baseline and 12-month follow-up | 44 (46%) | 45 (47%) | 1.03 (0.59 to 1.77) | 0.927 |
| Improvement in the AOU between baseline and 12-month follow-up | 10 (10%) | 12 (12%) | 1.25 (0.44 to 3.56) | 0.678 |
| **Relaxed per-protocol analysis** *(n = 261, clusters = 43)* | *N=123* | *N=138* | | |
| Improvement in the MAI score between baseline and 12-month follow-up | 47 (39%) | 58 (42%) | 1.18 (0.67 to 2.07) | 0.568 |
| Improvement in the AOU between baseline and 12-month follow-up | 17 (14%) | 20 (14%) | 1.09 (0.48 to 2.47) | 0.836 |

Acronyms: AOU=Assessment of Underutilization, CI=Confidence Interval, MAI=Medication Appropriateness Index, OR=Odds Ratio. | Calculated using multiple imputed data.

### eTable 9. Secondary outcomes: Per-protocol analysis

| | *Control (n=163)* | *Intervention (n=160)* | | *p-value* |
|---|---|---|---|---|
| **Medication-related secondary outcomes** | | | | |
| | *n (%)* | | *OR (95% CI)* | |
| Improvement in the MAI score between baseline and 6-month follow-up | 42 (44%) | 47 (48%) | 1.23 (0.62 to 2.44) | 0.561 |

| | | | | |
|---|---|---|---|---|
| Improvement in the AOU between baseline and 6-month follow-up | 14 (14%) | 11 (11%) | 0.80 (0.29 to 2.17) | 0.658 |
| | *Mean (95% CI)* | | *IRR (95% CI)* [4] | |
| MAI total score at 6-month follow-up | 23 (17; 29) | 27 (22; 33) | 1.87 (1.19 to 2.95) | 0.007 |
| MAI total score at 12-month follow-up | 24 (18; 31) | 29 (23; 34) | 1.50 (0.94 to 2.38) | 0.088 |
| Total number of prescribing omissions at 6-month follow-up as measured by the AOU | 1 (1; 1) | 1 (1; 1) | 1.07 (0.77 to 1.48) | 0.691 |
| Total number of prescribing omissions at 12-month follow-up as measured by the AOU | 1 (1; 1) | 1 (1; 1) | 0.82 (0.60 to 1.12) | 0.216 |
| | *Mean (95% CI)* | | *Mean difference (95% CI)* [4] | |
| Number of medications at 6-month follow-up | 8 (7; 9) | 8 (8; 9) | 0.51 (-0.30 to 1.32) | 0.219 |
| Number of medications at 12-month follow-up | 8 (8; 9) | 9 (8; 10) | 0.55 (-0.51 to 1.61) | 0.312 |
| **Patient-reported secondary outcomes** | | | | |
| | *Mean (95% CI)* | | *IRR (95% CI)* [4] | |
| Number of falls at 6-month follow-up | 0 (0; 0) | 0 (0; 0) | 0.67 (0.28 to 1.60) | 0.362 |
| Number of falls at 12-month follow-up | 0 (0; 0) | 0 (0; 0) | 0.95 (0.45 to 1.98) | 0.885 |
| | *Mean (95% CI)* | | *OR (95% CI)* | |
| Any fracture(s) between baseline and 6-month follow-up[3] | 4 (5%) | 1 (1%) | 0.22 (0.02 to 1.91) | 0.168 |
| Any fracture(s) between baseline and 12-month follow-up [3] | 2 (2%) | 2 (2%) | 0.89 (0.13 to 6.11) | 0.904 |
| *Quality of life* | | | | |
| | *Mean (95% CI)* | | *Mean difference (95% CI)* [4] | |
| EQ-5D-5L utilities at 6-month follow-up[1] (inverse predicted utility – IPU) | 0 (0 to 0) | 0 (0 to 0) | -0.05 (-0.11 to 0.01) | 0.067 |
| EQ-5D-5L utilities at 12-month follow-up[1] (inverse predicted utility – IPU) | 0 (0 to 0) | 0 (0 to 0) | 0.00 (-0.05 to 0.04) | 0.904 |
| Visual analogue scale at 6-month follow-up[2] | 70 (67; 74) | 72 (69; 76) | 2.67 (-1.79 to 7.14) | 0.240 |
| Visual analogue scale at 12-month follow-up[2] | 72 (69; 75) | 73 (70; 76) | 1.82 (-2.21 to 5.84) | 0.376 |

Acronyms: AOU=Assessment of Underutilization, CI=Confidence Interval, IRR=Incident Rate Ratio, MAI=Medication Appropriateness Index, MD=Mean difference, OR=Odds Ratio. This table is based on multiple imputed data and the strict per-protocol set of patients. | [1]Calculated based on the German Value Set for the EQ-5D-5L by Ludwig et al. PharmacoEconomics. 2018. | [2]Measured by visual analogue scale of the European Quality of Life-5 Dimensions questionnaire (EQ-VAS). Values range from 0 to 100 with higher values indicating a higher quality of life. | [3]Due to the low number of fractures a binary variable (any fracture yes/no) was considered. | [4]Adjusted for baseline characteristics.

### eTable 10. Additional secondary outcomes: Comparison between the intervention and control group

| | *Control (n=163)* | *Intervention (n=160)* | | *p-value* |
|---|---|---|---|---|
| **Additional secondary outcomes** | | | | |
| | *Number of events (%)*[3] | | *IRR (95% CI)*[4] | |
| Averaged MAI at 6-month follow-up[1] | 140 (20) | 143 (20) | 1.42 (0.95 to 2.12) | 0.084 |
| Averaged MAI at 12-month follow-up[1] | 141 (19) | 149 (14) | 1.29 (0.84 to 1.98) | 0.250 |
| Averaged number of prescribing omissions at the 6-month follow-up[2] | 135 (25) | 133 (30) | 0.92 (0.65 to 1.29) | 0.625 |
| Averaged number of prescribing omissions at the 12-month follow-up[2] | 133 (27) | 138 (25) | 0.74 (0.53 to 1.04) | 0.0.81 |

Acronyms: AOU=Assessment of Underutilization, CI=Confidence Interval, IRR=Incident Rate Ratio, MAI=Medication Appropriateness Index. This table is based on multiple imputed data. | [1]The total MAI score divided by the total number of chronic medications. | [2]The total number of prescribing omissions divided by the total number of chronic conditions. | [3]Number of patients with data (with multiple imputations). | [4]Adjusted for baseline characteristics.

### eTable 11. Descriptive secondary outcomes.

| | **Control group (n = 163)** | | **Intervention group (n = 160)** | |
|---|---|---|---|---|
| | *Missing n (%)* | *median [IQR]; mean (sd) or n (%)* | *Missing n (%)* | *median [IQR]; mean (sd) or n (%)* |
| *Falls* | | | | |
| Number of falls in the last 6 months (baseline) | 10 (6%) | | 5 (3%) | |
| 0 | | 128 (79%) | | 125 (78%) |
| 1 | | 19 (12%) | | 25 (16%) |
| 2 | | 4 (2.5%) | | 3 (1.9%) |
| >2 | | 2 (1.2%) | | 2 (1.3%) |
| Number of falls in the last 6 months (6-month follow-up) | 17 (10%) | | 18 (11%) | |
| 0 | | 121 (74%) | | 114 (71%) |
| 1 | | 20 (12%) | | 19 (12%) |
| 2 | | 2 (1.2%) | | 8 (5%) |
| >2 | | 3 (1.8%) | | 1 (0.6%) |
| Number of falls in the last 6 months (12-month follow-up) | 20 (12%) | | 20 (13%) | |
| 0 | | 118 (72%) | | 119 (74%) |
| 1 | | 21 (13%) | | 17 (11%) |
| 2 | | 3 (1.8%) | | 2 (1.3%) |
| >2 | | 1 (0.6%) | | 2 (1.3%) |
| *Fractures* | | | | |
| Number of fractures in the last 6 months (baseline) | 8 (5%) | | 4 (3%) | |
| 0 | | 152 (93%) | | 152 (95%) |
| 1 | | 3 (1.8%) | | 4 (2.5%) |
| 2 | | 0 (0%) | | 0 (0%) |

| | | | | |
|---|---|---|---|---|
| >2 | | 0 (0%) | | 0 (0%) |
| Number of fractures in the last 6 months (6-month follow-up) | 18 (11%) | | 18 (11%) | |
| 0 | | 141 (87%) | | 139 (87%) |
| 1 | | 2 (1.2%) | | 3 (1.9%) |
| 2 | | 0 (0%) | | 0 (0%) |
| >2 | | 2 (1.2%) | | 0 (0%) |
| Number of fractures in the last 6 months (12-month follow-up) | 20 (12%) | | 19 (12%) | |
| 0 | | 141 (87%) | | 138 (86%) |
| 1 | | 2 (1.2%) | | 3 (1.9%) |
| 2 | | 0 (0%) | | 0 (0%) |
| >2 | | 0 (0%) | | 0 (0%) |
| *Quality of life*[1] | | | | |
| EQ-5D-5L – Mobility (baseline) | 16 (10%) | | 8 (5%) | |
| No problems | | 59 (36%) | | 69 (43%) |
| Slight problems | | 35 (21%) | | 34 (21%) |
| Moderate problems | | 32 (20%) | | 28 (17%) |
| Severe problem | | 20 (12%) | | 21 (13%) |
| Unable | | 1 (0.61%) | | 0 (0.00%) |
| EQ-5D-5L – Mobility (6-month follow-up) | 26 (16%) | | 22 (14%) | |
| No problems | | 50 (31%) | | 58 (36%) |
| Slight problems | | 36 (22%) | | 37 (23%) |
| Moderate problems | | 35 (21%) | | 33 (21%) |
| Severe problem | | 15 (9.2%) | | 8 (5.0%) |
| Unable | | 1 (0.61%) | | 2 (1.3%) |
| EQ-5D-5L – Mobility (12-month follow-up) | 25 (15%) | | 25 (16%) | |
| No problems | | 63 (39%) | | 75 (47%) |
| Slight problems | | 37 (23%) | | 37 (23%) |
| Moderate problems | | 32 (20%) | | 18 (11%) |
| Severe problem | | 5 (3.1%) | | 5 (3.1%) |
| Unable | | 1 (0.61%) | | 0 (0.00%) |
| EQ-5D-5L – Selfcare (baseline) | 16 (10%) | | 8 (5%) | |
| No problems | | 124 (76%) | | 134 (84%) |
| Slight problems | | 13 (8.0%) | | 11 (6.9%) |
| Moderate problems | | 7 (4.3%) | | 4 (2.5%) |
| Severe problem | | 2 (1.2%) | | 2 (1.3%) |
| Unable | | 1 (0.61%) | | 1 (0.63%) |
| EQ-5D-5L – Selfcare (6-month follow-up) | 26 (16%) | | 22 (14%) | |
| No problems | | 119 (73%) | | 118 (74%) |
| Slight problems | | 10 (6.1%) | | 15 (9.4%) |
| Moderate problems | | 6 (3.7%) | | 3 (1.9%) |
| Severe problem | | 1 (0.61%) | | 1 (0.63%) |
| Unable | | 1 (0.61%) | | 1 (0.63%) |
| EQ-5D-5L – Selfcare (12-month follow-up) | 25 (15%) | | 25 (16%) | |
| No problems | | 120 (74%) | | 116 (73%) |

| | | | | |
|---|---|---|---|---|
| Slight problems | | 13 (8.0%) | | 13 (8.1%) |
| Moderate problems | | 3 (1.8%) | | 6 (3.8%) |
| Severe problem | | 2 (1.2%) | | 0 (0.00%) |
| Unable | | 0 (0.00%) | | 0 (0.00%) |
| EQ-5D-5L – Activities (baseline) | 16 (10%) | | 8 (5%) | |
| No problems | | 99 (61%) | | 104 (65%) |
| Slight problems | | 19 (12%) | | 16 (10%) |
| Moderate problems | | 24 (15%) | | 22 (14%) |
| Severe problem | | 3 (1.8%) | | 8 (5.0%) |
| Unable | | 2 (1.2%) | | 2 (1.3%) |
| EQ-5D-5L – Activities (6-month follow-up) | 26 (16%) | | 22 (14%) | |
| No problems | | 75 (46%) | | 82 (51%) |
| Slight problems | | 34 (21%) | | 31 (19%) |
| Moderate problems | | 18 (11%) | | 20 (13%) |
| Severe problem | | 8 (4.9%) | | 4 (2.5%) |
| Unable | | 2 (1.2%) | | 1 (0.63%) |
| EQ-5D-5L – Activities (12-month follow-up) | 25 (15%) | | 26 (16%) | |
| No problems | | 90 (55%) | | 96 (60%) |
| Slight problems | | 25 (15%) | | 25 (16%) |
| Moderate problems | | 18 (11%) | | 11 (6.9%) |
| Severe problem | | 5 (3.1%) | | 2 (1.3%) |
| Unable | | 0 (0.00%) | | 0 (0.00%) |
| EQ-5D-5L – Pain or discomfort (baseline) | 16 (10%) | | 8 (5%) | |
| No problems | | 61 (37%) | | 60 (38%) |
| Slight problems | | 31 (19%) | | 33 (21%) |
| Moderate problems | | 41 (25%) | | 43 (27%) |
| Severe problem | | 14 (8.6%) | | 13 (8.1%) |
| Unable | | 0 (0.00%) | | 3 (1.9%) |
| EQ-5D-5L – Pain or discomfort (6-month follow-up) | 26 (16%) | | 22 (14%) | |
| No problems | | 25 (15%) | | 30 (19%) |
| Slight problems | | 51 (31%) | | 47 (29%) |
| Moderate problems | | 43 (26%) | | 50 (31%) |
| Severe problem | | 17 (10%) | | 11 (6.9%) |
| Unable | | 1 (0.61%) | | 0 (0.00%) |
| EQ-5D-5L – Pain or discomfort (12-month follow-up) | 25 (15%) | | 25 (16%) | |
| No problems | | 40 (25%) | | 41 (26%) |
| Slight problems | | 53 (33%) | | 46 (29%) |
| Moderate problems | | 37 (23%) | | 37 (23%) |
| Severe problem | | 8 (4.9%) | | 11 (6.9%) |
| Unable | | 0 (0.00%) | | 0 (0.00%) |
| EQ-5D-5L – Anxiety (baseline) | 17 (10%) | | 8 (5%) | |
| No problems | | 95 (58%) | | 99 (62%) |
| Slight problems | | 29 (18%) | | 33 (21%) |
| Moderate problems | | 19 (12%) | | 18 (11%) |
| Severe problem | | 3 (1.8%) | | 2 (1.3%) |
| Unable | | 0 (0.00%) | | 0 (0.00%) |
| EQ-5D-5L – Anxiety (6-month follow-up) | 26 (16%) | | 22 (14%) | |

| | | | | |
|---|---|---|---|---|
| No problems | | 77 (47%) | | 84 (52%) |
| Slight problems | | 40 (25%) | | 29 (18%) |
| Moderate problems | | 15 (9.2%) | | 22 (14%) |
| Severe problem | | 4 (2.5%) | | 3 (1.9%) |
| Unable | | 1 (0.61%) | | 0 (0.00%) |
| EQ-5D-5L – Anxiety (12-month follow-up) | 25 (15%) | | 25 (16%) | |
| No problems | | 95 (58%) | | 88 (55%) |
| Slight problems | | 30 (18%) | | 34 (21%) |
| Moderate problems | | 12 (7.4%) | | 12 (7.5%) |
| Severe problem | | 1 (0.61%) | | 1 (0.63%) |
| Unable | | 0 (0.00%) | | 0 (0.00%) |
| *EQ-5D-5L utilities*[2] | | | | |
| Baseline | 17 (10%) | 0.89 [0.81, 0.94]; 0.83 (0.18) | 8 (5%) | 0.89 [0.77, 0.97]; 0.83 (0.20) |
| 6-month follow-up | 26 (16%) | 0.86 [0.77, 0.92]; 0.79 (0.22) | 22 (14%) | 0.87 [0.80, 0.94]; 0.83 (0.18) |
| 12-month follow-up | 25 (15%) | 0.91 [0.82, 0.94]; 0.86 (0.14) | 26 (16%) | 0.91 [0.83, 0.94]; 0.87 (0.15) |
| *EQ-5D-5L - Visual analogue scale (VAS)*[3] | | | | |
| Baseline VAS | 17 (10%) | 70 [60, 80]; 70 (17) | 10 (6%) | 75 [60, 80]; 71 (17) |
| 6-month follow-up VAS | 26 (16%) | 75 [60, 85]; 71 (17) | 22 (14%) | 75 [60, 80]; 72 (16) |
| 12-month follow-up VAS | 25 (15%) | 75 [60, 80]; 73 (15) | 27 (17%) | 75 [60, 85]; 73 (15) |

Acronyms: IQR=inter quartile range, SD=standard deviation, MAI=Medication Appropriateness Index, AOU=Assessment of underutilization, VAS=Visual Analogue Scale. | This table is based on the raw data. | [1]Quality of Life was assessed using the EQ-5D-5L questionnaire of the EuroQol group. | [2]Calculated based on the German Value Set for the EQ-5D-5L by Ludwig et al. PharmacoEconomics. 2018. | [3]Measured by visual analogue scale of the European Quality of Life-5 Dimensions questionnaire (EQ-VAS). Values range from 0 to 100 with higher values indicating a higher quality of life.
